# Prevalence of SARS-CoV-2 antibodies in France: results from nationwide serological surveillance

**DOI:** 10.1101/2020.10.20.20213116

**Authors:** Stéphane Le Vu, Gabrielle Jones, François Anna, Thierry Rose, Jean-Baptiste Richard, Sibylle Bernard-Stoecklin, Sophie Goyard, Caroline Demeret, Olivier Helynck, Corinne Robin, Virgile Monnet, Louise Perrin de Facci, Marie-Noelle Ungeheuer, Lucie Léon, Yvonnick Guillois, Laurent Filleul, Pierre Charneau, Daniel Lévy-Bruhl, Sylvie van der Werf, Harold Noel

## Abstract

**Background:** Assessment of cumulative incidence of SARS-CoV-2 infections is critical for monitoring the course and the extent of the epidemic. As asymptomatic or mild cases were typically not captured by surveillance data in France, we implemented nationwide serological surveillance. We present estimates for prevalence of anti-SARS-CoV-2 antibodies in the French population and the proportion of infected individuals who developed potentially protective neutralizing antibodies throughout the first epidemic wave.

**Methods:** We performed serial cross-sectional sampling of residual sera over three periods: prior to (9-15 March), during (6-12 April) and following (11-17 May) a nationwide lockdown. Each sample was tested for anti-SARS-CoV-2 IgG antibodies targeting the Nucleoprotein and Spike using two Luciferase-Linked ImmunoSorbent Assays, and for neutralising antibodies using a pseudo-neutralisation assay. We fitted a general linear mixed model of seropositivity in a Bayesian framework to derive prevalence estimates stratified by age, sex and region.

**Findings:** In total, sera from 11 021 individuals were analysed. Nationwide seroprevalence of SARS-CoV-2 antibodies was estimated at 0.41% [0.05−0.88] mid-March, 4.14% [3.31−4.99] mid-April and 4.93% [4.02−5.89] mid-May. Approximately 70% of seropositive individuals had detectable neutralising antibodies. Seroprevalence was higher in regions where circulation occurred earlier and was more intense. Seroprevalence was lowest in children under 10 years of age (2.72% [1.10−4.87]).

**Interpretation:** Seroprevalence estimates confirm that the nationwide lockdown substantially curbed transmission and that the vast majority of the French population remains susceptible to SARS-CoV-2. Low seroprevalence in school age children suggests limited susceptibility and/or transmissibility in this age group. Our results show a clear picture of the progression of the first epidemic wave and provide a framework to inform the ongoing public health response as viral transmission is picking up again in France and globally.

**Funding:** Santé publique France.

## Introduction

After the first case of SARS-CoV-2 infection was reported in France on 24 January 2020, authorities largely relied on confirmed case counts to monitor the unfolding epidemic.^1^ Case-based surveillance focused primarily on symptomatic patients or those with severe disease and access to biological confirmation was initially limited. The surge in COVID-19 hospitalisations and deaths led the French authorities to implement a general lockdown from 17 March to 11 May 2020.

It is now clear that a substantial fraction of infected individuals develop mild symptoms or even remain asymptomatic.^2–5^ For this reason, the actual proportion of the French population infected during the first epidemic wave remains elusive. Prevalence of previous or current infections is critical to understanding the course and extent of the epidemic. Since a serological response is likely to take place in all SARS-CoV-2 infected individuals, the corresponding serological markers should persist for at least some time. Accordingly, prevalence of SARS-CoV-2 antibodies can assess cumulative population incidence. Such an assessment can be obtained from seroepidemiological studies, provided that the antibody detection method is accurate enough, even in a low prevalence context, and that the results from the study sample can reasonably be extrapolated to the population. In addition, such studies can measure the proportion of infected individuals who developed neutralising and potentially protective antibodies which is particularly important in the absence of a vaccine.^6^ To the best of our knowledge, few seroprevalence studies have included detection of SARS-CoV-2 neutralising antibodies, and none at a national level.^2,7–10^

To estimate the fraction of the French population infected with SARS-CoV-2 over time as well as the proportion of individuals having developed neutralising antibodies, we implemented serological surveillance based on serial cross-sectional sampling of residual sera obtained from clinical laboratories. Here we present nationwide estimates of seroprevalence in the French population, as well as estimates stratified by age, sex and region, from three collection periods prior to, during and following the lockdown.

## Methods

### Design and population

Serological surveillance used repeated cross-sectional sampling of residual sera obtained from the biobanks of two of the largest centralizing laboratories in France covering all regions and accounting for 90% of the market share in specialty clinical diagnostic testing (Unpublished data). Residual sera included specimens from individuals of all ages undergoing routine diagnosis and monitoring in all medical specialties (biochemistry, immunology, allergy…) except infectious diseases and obstetrics.

### Sample selection and preparation

Specimens were collected over three one-week periods: prior to (9-15 March 2020), during (6-12 April 2020) and following (11-17 May 2020) the nationwide lockdown. To obtain results by subgroups and enough precision, we randomly sampled available sera at the biobanks. Sampling was stratified by sex, 10-year age groups (0-9 years to ≥80 years) and region. Due to the limited number of sera available for French overseas departments (Guadeloupe, Martinique, Mayotte, French Guiana, La Réunion), all available sera were included. Relying on early modelling of the COVID-19 epidemic, which estimated an expected prevalence of 3% as of 28 March 2020, we calculated a target sample size of 3,500 per collection period, with a margin of error of 0.55%.^11^ After selection, blood samples were centrifuged and sera were transferred on 96-well microplates then frozen at −20°C before transport.

### SARS-COV-2 antibody testing

All serological analyses were conducted with the National Reference Centre (NRC) for Respiratory Infection Viruses including Influenza at the Institut Pasteur in Paris. Three novel serological assays were developed: two LuLISAs (Luciferase-Linked ImmunoSorbent Assay), detecting the nucleoprotein (LuLISA N) and spike (LuLISA S) protein of SARS-CoV-2 respectively, and a pseudo-neutralisation assay (PNT).^12,13^ The two LuLISA assays are endowed with a wide dynamic range (4-log) and a high throughput capacity (2300 assays/h).^14^ In LuLISA, the presence of all four anti-N or anti-S IgG subtypes is revealed by the means of a unique alpaca anti-human VhH (single variable heavy chain antibody domain) coupled to a luciferase, the bioluminescent activity of which is then measured. Serum samples are considered positive when the relative light units per second (RLU/s) value is above the threshold determined for each of the LuLISA IgG/N and IgG/S assays from a pre-pandemic serum collection. The PNT mimics the SARS-CoV-2 entry step in HEK 293T cells stably expressing the human SARS-CoV-2 spike receptor ACE2 on their surface. It uses a lentiviral vector pseudo-typed with SARS-CoV-2 Spike protein, which penetrates cells in an ACE2-dependent manner, and consequently expresses a luciferase Firefly reporter. When the lentiviral Spike-mediated entry is blocked by potential serum neutralising antibodies, this leads to a reduced bioluminescence signal expressed as RLU/s and samples are considered positive with values below a threshold set as the mean minus 3-fold the standard deviation determined on a collection of pre-pandemic sera. This test makes it possible to estimate the prevalence of potentially neutralising anti-S antibodies, although the effective level of protection conferred by neutralising antibodies remains unclear.

### Assay calibration

Individual test characteristics were assessed using sets of pre-pandemic sera collected before 04/09/2019 and sera from hospitalised cases of COVID-19 confirmed by RT-PCR sampled at least 7 days past symptoms onset (Figure S1). In a context of low expected prevalence of infections, we set the thresholds to define a positive test result in order to obtain a 100% specificity to reduce the risk of false positives. This led to suboptimal sensitivities for each individual testing method, ranging from 85 % to 96 % (Table S1). Since individuals exposed to the SARS-CoV-2 virus do not undergo a single type of immune response, the results of three different but complementary serological tests provided a more precise assessment of the population exposure to the virus. We defined seroprevalence based on the proportion of individuals who tested positive for SARS-CoV-2 antibodies for at least one of the three tests. This combination led to a perfect classification for our set of reference samples (223 prepandemic subjects and 45 hospitalised confirmed cases of COVID-19) (Table S1).

### Statistical analysis

We inferred probability of SARS-CoV-2 seropositivity in the population using (i) the proportion of specimens having at least one positive result among the three tests (ii) the calibration study to account for biological test performance and (iii) the poststratification variables to account for demographic and geographic differences between the sample and general population.

We fitted a general linear mixed model of seropositivity explained by gender, age, region, and the collection period in a Bayesian framework.^15^ Actual seroprevalence was derived from the frequency of positive tests, using estimates and associated uncertainties for sensitivity and specificity obtained from the calibration study. The resulting probabilities of seropositivity in each stratum were used to derive post-stratified estimates for the total population and by subgroups, using national census population counts stratified by gender, 10-year age bands and region.^16^ Using estimates of regression model coefficients, we calculated risk of infection relative to a reference category for each covariate.

The model was developed using RStan and all data processing used R.^17,18^ Code is available as online supplementary material. Estimates are reported as mean of the posterior probability distributions over 10^4^ iterations and their uncertainty intervals by the 2.5th and 97.5th percentiles of the distributions. Further details of the model are available in the supplementary materials.

## Ethical considerations

Authorization for conservation and preparation of elements of the human body for scientific use was granted to the two biobanks by the bioethics committee from General Board for Research and Innovation (DGRI) of French Ministry of Higher Education and Research (approvals N°AC-2015-2418 and AC-2018-3329). Information regarding secondary use of de-identified residual sera for approved research studies was systematically displayed and orally communicated at the primary clinical laboratories. The Ethics Committee (Comité de Protection des Personnes Ile-de-France VI, CHU Pitié-Salpétrière Hospital, Paris, France) waived the need for ethical approval for the collection, analysis and publication of the retrospectively obtained and anonymized specimens and data for this study. This work was carried out following regulations of the French Public Health Code (articles L. 1413-7 and L. 1413-8) and the French Commission for Data Protection (CNIL).

## Results

A total of 9 184 residual sera for Metropolitan France were randomly selected from available sera at the three collection periods (3 221 samples from 9-15 March, 3 084 samples from 6-12 April, and 2 879 samples from 11-17 May). For the French overseas departments, 613, 511 and 713 samples were included respectively for the three collection periods (we excluded Mayotte Island from the analysis due to an insufficient number of available samples). The age, sex and regional distribution of the sample population is shown in Table S2.

Nationwide seroprevalence of SARS-CoV-2 infections increased from 0.41% [0.05−0.88] to 4.14% [3.31−4.99] and 4.93% [4.02−5.89] between 15 March, 12 April and 17 May, corresponding to 3 292 000 [2 685 000−3 934 000] people having been infected as of 17 May (Table S4). Note that when taking into account the inherent delay between infection and IgG-mediated antibody responses, this estimate provides the number of infections which occurred approximately two weeks prior to the collection periods.^19^ The prevalence of pseudo-neutralising antibodies for SARS-CoV-2 S-protein rose from 0.06% [0.00 −0.17] to 3.33% [2.66-4.07] over the same period (Table 1). The raw proportions of positive sera for each individual test are detailed in Table S3. Seroprevalence increased significantly between March and April, with a ten-fold increase in relative risk, but plateaued from April to May (Figure 1).

**Table 1:**
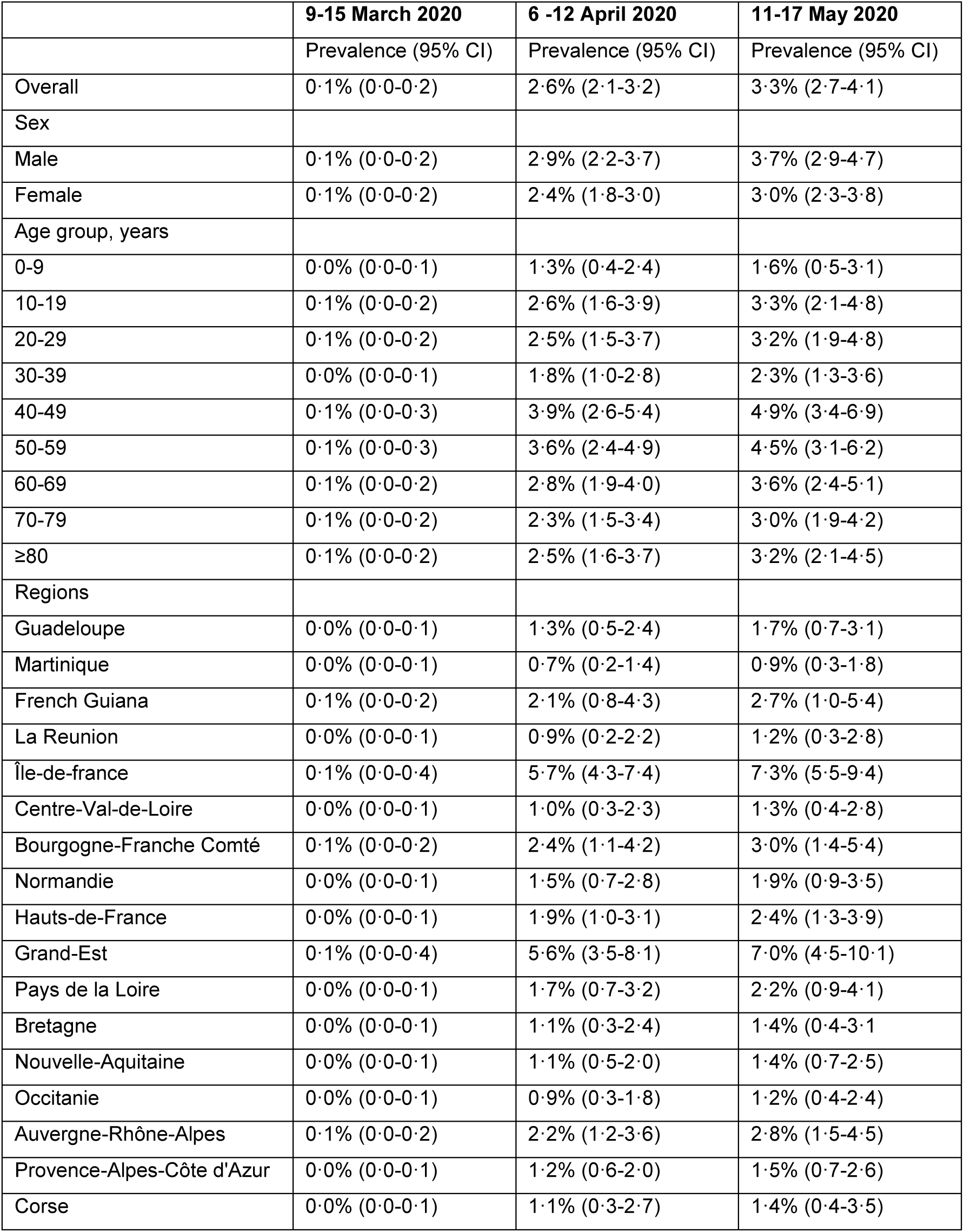
Estimated prevalence of SARS-CoV-2 neutralising antibodies in the French population from March-May 2020.

**Figure 1:**
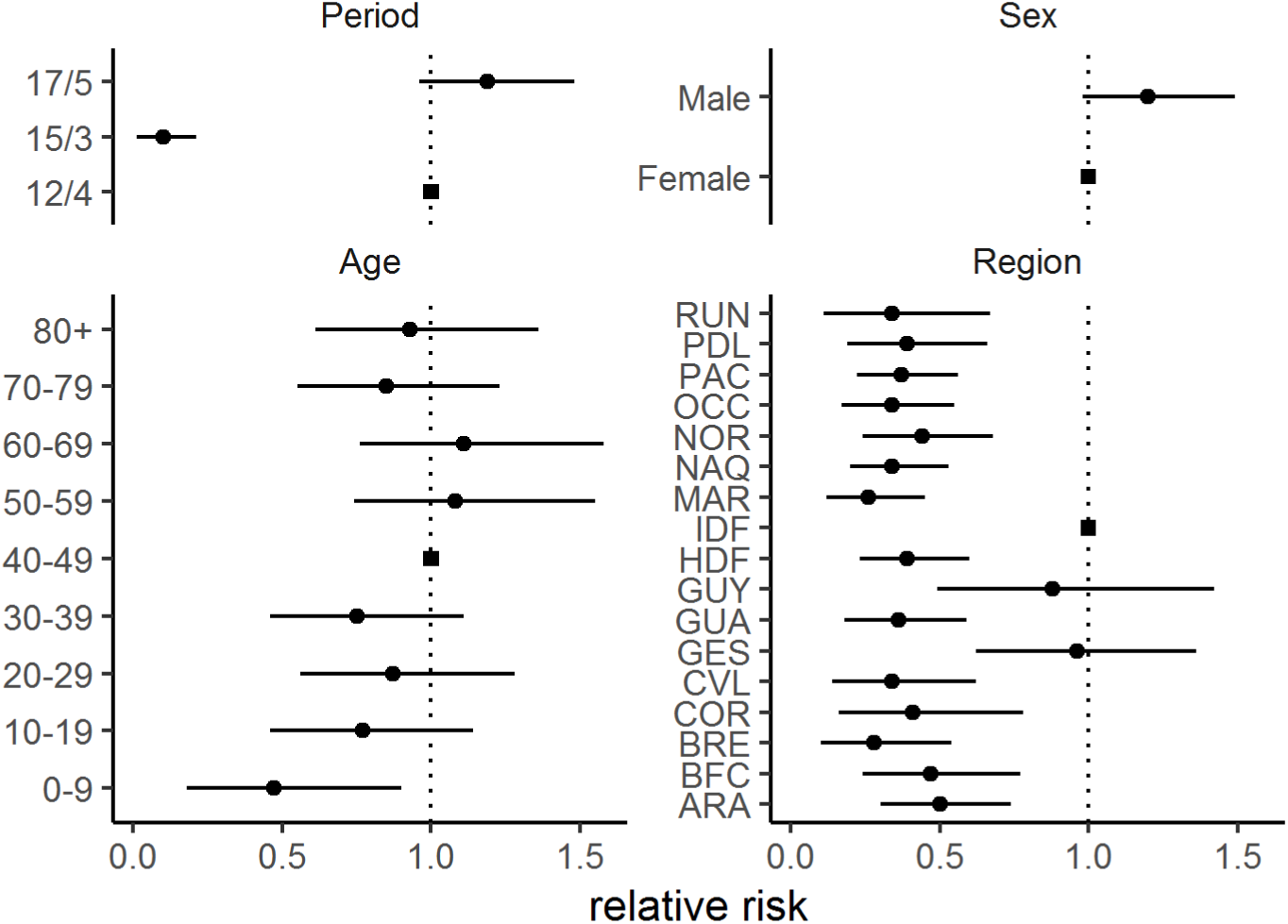
Estimated relative risks of seroprevalence by collection period, sex, age and region. Reference categories are indicated by a square. Bars represent 95% uncertainty interval of relative risk estimate over 104 iterations. Collection periods are indicated by last day of each week (9-15 March 2020, 6-12 April 2020, 11-17 May 2020). Regions: GUA = Guadeloupe, MAR = Martinique, GUY = French Guiana, RUN = La Réunion, IDF = Île-de-France, CVL = Centre-Val-de-Loire, BFC = Bourgogne-Franche Comté, NOR = Normandie, HDF = Hauts-de-France, GES = Grand-Est, PDL = Pays de la Loire, BRE = Bretagne, NAQ = Nouvelle-Aquitaine, OCC = Occitanie, ARA = Auvergne-Rhône-Alpes, PAC = Provence-Alpes-Côtes d’Azur, COR = Corse.

No significant difference in seroprevalence was observed based on sex (5.37% [4.27−6.55] for men and 4.51% [3.57−5.54] for women at 17 May). At the same time point, 3.70% [2.87−4.65] of male and 2.98% [2.26−3.81] of female individuals had detectable pseudo-neutralising antibodies.

From mid-March to mid-May, the prevalence of infections increased markedly in all age groups (Figure 2). As of 17 May, one week after the end of lockdown, the prevalence was highest among the 50−59 year and 60-69 years olds (6.06% [4.43−8.04], and 6.04% [4.40-8.06] respectively), and lowest in children under 10 years of age (2.72% [1.10−4.87]). Prevalence of pseudo-neutralising antibodies followed similar trends and varied according to age from 1.59% [0.52−3.13] in children under 10 years old to 4.92% [3.36−6.89] in 40-49 year olds, then decreasing in older aged groups (Table 1).

**Figure 2:**
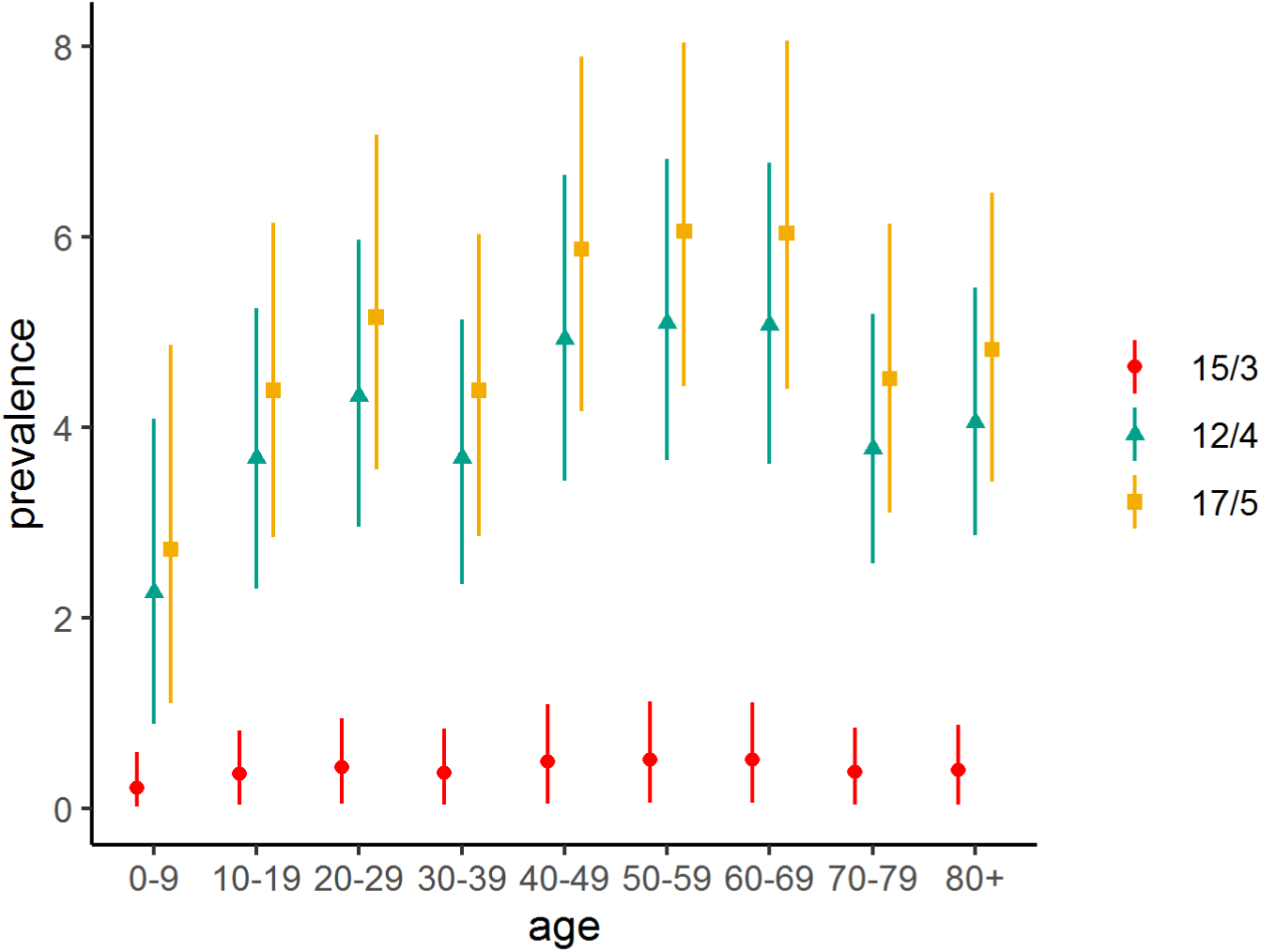
Estimated prevalence of SARS-CoV-2 antibodies in the French population by age group and collection period. Collection periods are indicated by last day of each week (9-15 March 2020, 6-12 April 2020, 11-17 May 2020). Bars represent 95% uncertainty interval of prevalence estimate over 104 iterations.

Regional seroprevalence was highest in Île-de-France which includes Paris (8.82% [6.90−11.01]) and Grand-Est (8.56% [5.83−11.82]) in the week after lockdown was lifted and varied from 2.40% to 4.44% in the remaining Metropolitan regions with a clear East-West gradient (Figure 3). Seroprevalence in the four overseas regions ranged from 2.40% [1.18−3.93] in Martinique to 7.14% [3.96−11.50] in French Guiana (Figure 3). Regional prevalence estimates of pseudo-neutralising antibodies followed similar trends and were highest in Île-de-France (7.25% [5.51−9.36]) and in Grand-Est (7.03% [4.48−10.06]).Estimates ranged from 1.20% to 3.03% for the remaining Metropolitan regions and from 0.86% to 2.66% in overseas regions (Table S4).

**Figure 3:**
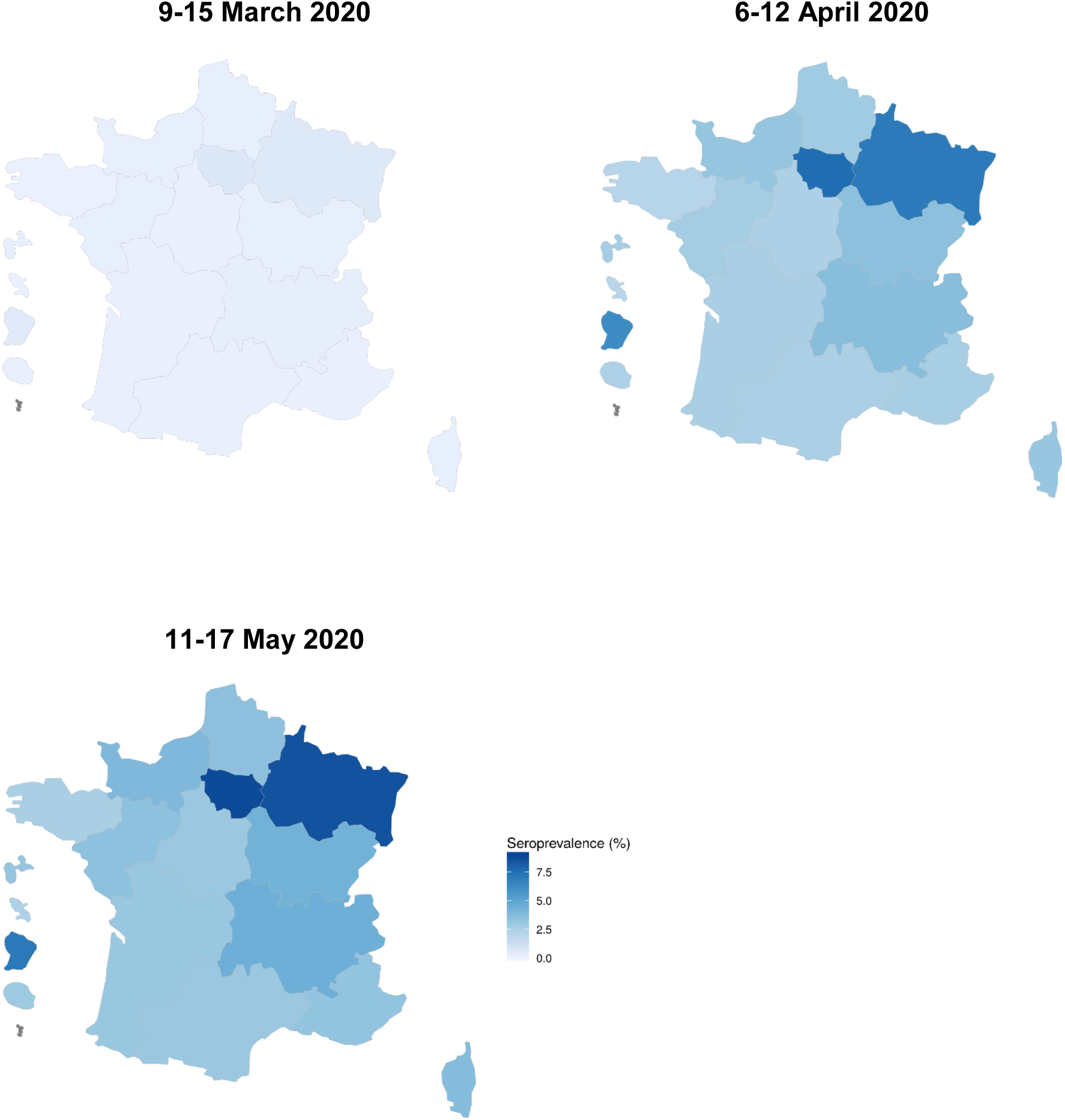
Estimated prevalence of SARS-CoV-2 antibodies in the French population by region.

## Discussion

Nationwide serological surveillance in France shows that following the first wave of the COVID-19 epidemic, seroprevalence remained low, with about 5% of the population having developed a detectable humoral response to the virus. Our results are within the same order of magnitude as studies carried out at comparable epidemic stages in Europe.^4,20–22^ Estimates at multiple points of the French epidemic show a sharp increase between the first two collection periods, immediately preceding and during the generalized lockdown, followed by little progression observed at the final collection period just after lockdown ended. This confirms its substantial impact in almost halting community transmission.

One of the primary strengths of our study is the inclusion of individuals of all ages, notably children under 10 years old. Understanding how school-aged children are susceptible to infection remains of particular importance in the face of continuing challenges for public health decisions about school settings. Seroprevalence was lowest in school-aged children suggesting limited susceptibility and/or transmissibility in this age group. A previous cohort study in France concluded that primary school aged children were poor drivers of SARS-CoV-2 transmission amongst themselves or to teachers.^23^

As expected, regional results show significantly higher seroprevalence where circulation occurred earlier and was more intense, notably in Île-de-France and Grand-Est. A large religious gathering in early March in the Grand-Est region triggered intense regional circulation of the virus and was responsible for secondary cases all over Metropolitan France and in French Guiana.^24^ Estimates for other French regions confirm widespread, but less intense SARS-CoV-2 circulation at the exit of lockdown.

To date, few seroprevalence studies have included detection of neutralising antibodies, which are theoretically correlated to protection.^2,3,7,10^ Importantly, seroprevalence of neutralising antibodies has not been estimated at a nationwide scale. As of 17 May, we find that approximately 70% of seropositive individuals had detectable pseudo-neutralising antibodies with large variation across age categories and regions. Several studies similarly reported that only a fraction of seropositive individuals had detectable levels of neutralising antibodies, this fraction being variable.^2,3,7–9^ This finding could be explained by differences in antibody kinetics with delayed appearance of neutralising antibodies.^12^

There are three additional factors which should be taken in account in the interpretation of our results. First, we set positive thresholds for our assays to a specificity of 100%. While ruling out the risk of false positives, this could preclude the detection of the lowest antibody levels. Especially, our in-house tests were calibrated on a series of confirmed, hospitalised, COVID-19 cases, which may have limited the assessment of sensitivity. As a result, and even though the model corrected for imperfect sensitivity, we may still be underestimating the proportion of individuals with mild or asymptomatic infections who may develop a weaker or more short-lived humoral response.^25–27^ Moreover, possible differential waning of antibody levels affecting mainly anti N and pseudo-neutralising antibodies, could also result in an underestimation of seroprevalence at a distance from the epidemic waves, but this should be negligible within our relatively short surveillance period.^12,28,29^ In order to facilitate the interpretation of SARS-CoV-2 infection seroprevalence as the pandemic progresses, further longitudinal serological studies documenting symptoms and immune response are essential. Finally, although the use of residual sera limits the risk of self-selection bias, it may introduce potential bias if individuals who required laboratory tests differ in terms of risk of infection from the general population. If the sampled individuals required routine monitoring for chronic health problems, they may have taken greater precautions and lowered their exposure to the virus, leading to underestimation of seroprevalence compared to the general population.

Although sample representativeness remains difficult to assess, our estimates are comparable to those reported from serological studies conducted in large pre-existing representative cohorts in Île-de France, Grand-Est and Nouvelle-Aquitaine.^2^ Additionally, when comparing our regional estimates and COVID-19 mortality rates, a surveillance indicator with a low susceptibility to reporting bias and which should correlate with population exposure, we find a strong correlation (r=0.81), with French Guiana largely influencing the overall coefficient (Figure S2). This discrepancy between virus circulation and mortality rate for French Guiana seems to be explained by the age structure of its infected population, skewed towards young ages.^30^ These assesments against external data suggest that using residual sera can be a robust and cost-effective approach for serological surveillance. The availability of residual sera made it possible to quickly implement sample collection early in the epidemic, providing a background seroprevalence estimate prior to the peak, and to observe epidemic dynamics throughout the first wave by including multiple collection periods. Our seroprevalence estimates, including the proportion of the population having produced pseudo-neutralising antibodies, confirmed that post-lockdown, the vast majority of the French population remained susceptible to SARS-CoV-2, even in regional hotspots. We find that a seroprevalence of at most 9% in certain regions yielded enough hospitalisations to overwhelm the healthcare system. Our results provide a critical understanding of the progression of the first epidemic wave and provide a framework to inform the ongoing public health response as viral transmission is picking up again in France and globally. Serological surveillance based on residual sera will continue to be used to provide timely seroprevalence estimates as the epidemic evolves and through the 2020-2021 winter season to monitor the progression of population level immunity and guide public health response.

## Data Availability

Data from the analysis can be made available upon
request to the corresponding author and might
require partial aggregation or downsampling to
protect patient privacy.

## Acknowledgements

We thank Christine Larsen, Bruno Coignard and Jean-Claude Desenclos (Santé publique France) for their helpful contibution at setting up the study; from Institut Pasteur, Isabelle Cailleau for support in the funding process, Hélène Munier for access to automate and supply management at the Unit of Chemistry and Biocatalysis, Yves Janin for provision of the luciferase substrate, Nicolas Escriou and Marion Gransagne for the design of the antigen expressing plasmids, Stephane Petres for production and supply of the N and S antigens, Philippe Souque for production of the lentiviral pseudo-types; Juliette Paireau (Institut Pasteur, Santé publique France), Rodolphe Thiébaut (Bordeaux Université) and Xavier de Lamballerie (Aix-Marseille Université) for valuable comments on first results. We also thank the teams from the Eurofins Biomnis Sample Library and Cerba Healthcare Division for contributing to sample collection.

## Author contributions

HN, GJ, J-BR, SBS, DLB, SvdW conceived and planned the study. VM, CR, M-NU, LPdF, OH contributed to sample collection. TR, FA, SG developed the assays and carried out the biological analyses. SLV, J-BR, HN, GJ designed and performed data analyses. HN, GJ, J-BR, SLV, SBS, DLB, TR, PC, FA, CD, SvdW, LL, YG, LF contributed to the interpretation of results. HN, GJ, SLV wrote the draft manuscript. All authors discussed the results and approved the final manuscript.

## Supplementary material

**Table S1:** Calibration of LuLISA N, LuLISA S et pseudo-neutralisation assays on prepandemic samples and confirmed COVID-19 cases.

|  | **Serological Assay** |  |  |  |
| --- | --- | --- | --- | --- |
|  | **LuLISA N** | **LuLISA S** | **PNT** | **LN\|LS\|PNT** |
| Readings threshold (RLU/s) | 39915 | 27821 | 14037 | as defined at left |
| Prepandemic samples | 523 | 232 | 223 | 223 |
| Confirmed COVID-19 cases | 309 | 45 | 99 | 45 |
| False positive | 0 | 0 | 0 | 0 |
| True positive | 266 | 43 | 84 | 45 |
| True negative | 523 | 232 | 223 | 223 |
| False negative | 43 | 2 | 15 | 0 |
| Specificity | 1.00 | 1.00 | 1.00 | 1.00 |
| Sensitivity | 0.86 | 0.96 | 0.85 | 1.00 |

LN=LuLISA N assay, LS=LuLISA S assay, PNT=Pseudo-neutralisation assay. Readings are expressed in relative light units per second (RLU/s). Confirmed COVID-19 cases were hospitalised patients with positive RT-PCR for SARS-CoV-2.

**Table S2:**
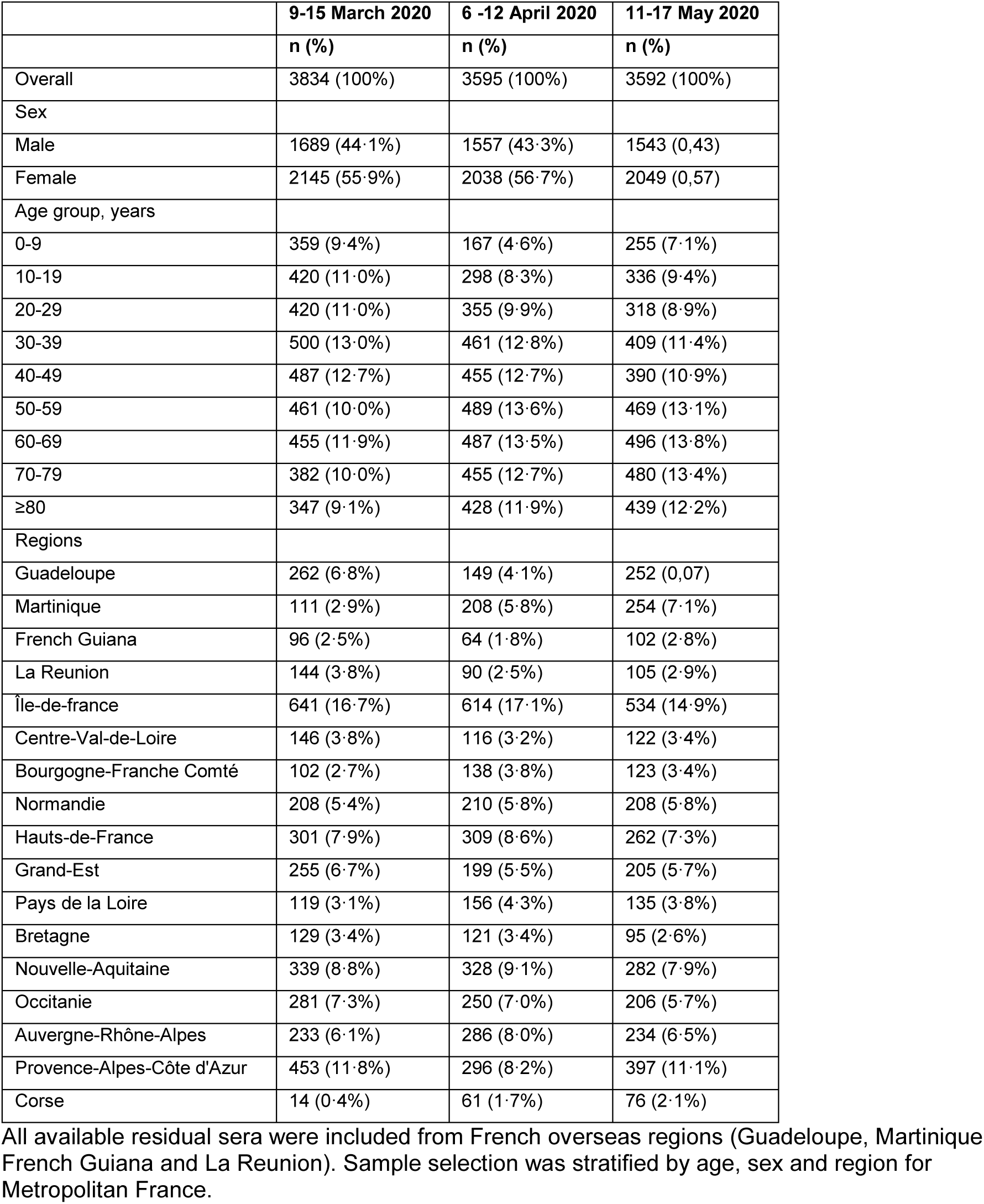
Distribution of sample specimens included by collection period.

**Table S3:**
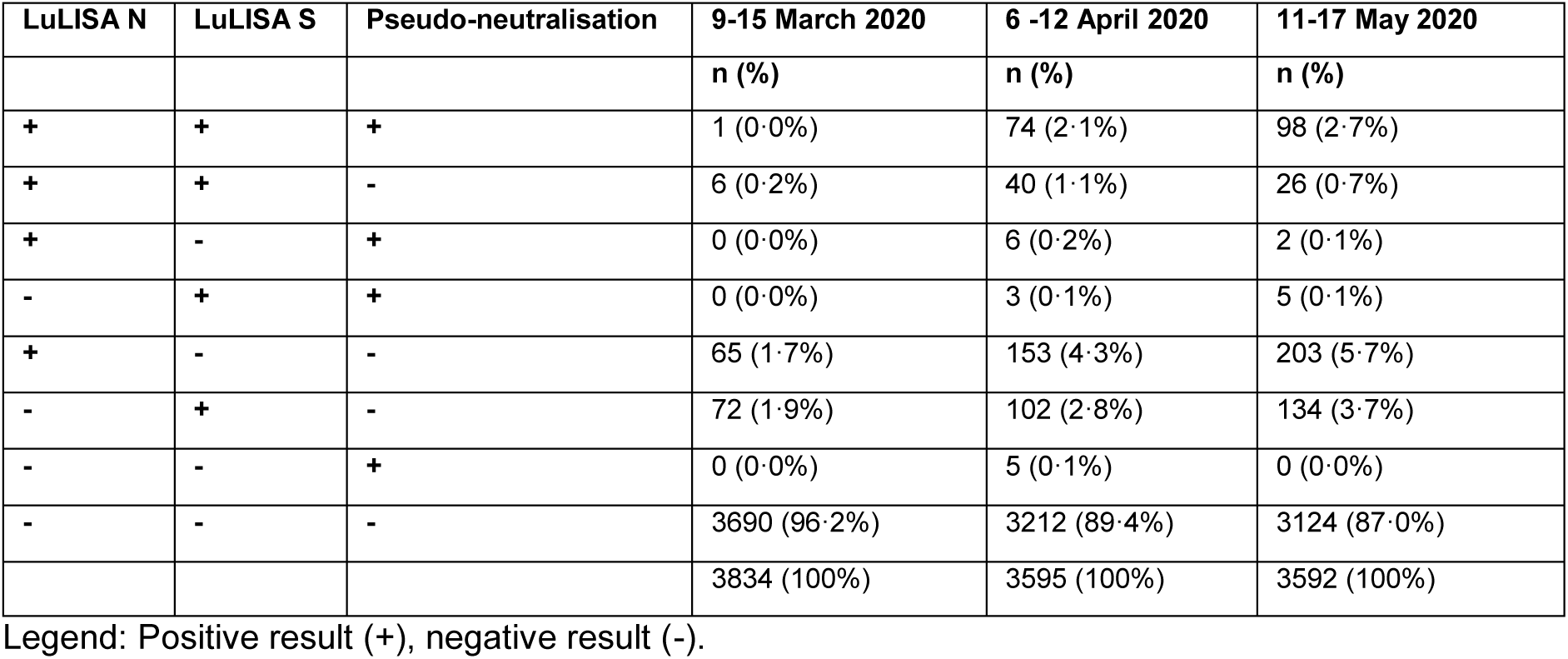
Combination of unadjusted results for the three serological assays by collection period.

**Table S4:** Estimated prevalence of SARS-CoV-2 antibodies in the French population from March-May 2020.

|  | **9-15 March 2020** | **6 -12 April 2020** | **11-17 May 2020** |
| --- | --- | --- | --- |
|  | Prevalence (95% CI) | Prevalence (95% CI) | Prevalence (95% CI) |
| Overall | 0.4% (0.1-0.9) | 4.1% (3.3-5.0) | 4.9% (4.0-5.9) |
| Sex |  |  |  |
| Male | 0.5% (0.1-1.0) | 4.5% (3.5-5.6) | 5.4% (4.3-6.6) |
| Female | 0.4% (0.0-0.8) | 3.8% (2.9-4.7) | 4.5% (3.6-5.5) |
| Age group, years |  |  |  |
| 0-9 | 0.2% (0.0-0.6) | 2.3% (0.9-4.1) | 2.7% (1.1-4.9) |
| 10-19 | 0.4% (0.0-0.8) | 3.7% (2.3-5.3) | 4.4% (2.9-6.2) |
| 20-29 | 0.4% (0.1-1.0) | 4.3% (3.0-6.0) | 5.2% (3.6-7.1) |
| 30-39 | 0.4% (0.0-0.8) | 3.7% (2.4-5.1) | 4.4% (2.9-6.0) |
| 40-49 | 0.5% (0.1-1.1) | 4.9% (3.4-6.7) | 5.9% (4.2-7.9) |
| 50-59 | 0.5% (0.1-1.1) | 5.1% (3.7-6.8) | 6.1% (4.4-8.0) |
| 60-69 | 0.5% (0.1-1.1) | 5.1% (3.6-6.8) | 6.0% (4.4-8.1) |
| 70-79 | 0.4% (0.0-0.9) | 3.8% (2.6-5.2) | 4.5% (3.1-6.1) |
| ≥80 | 0.4% (0.0-0.9) | 4.1% (2.9-5.5) | 4.8% (3.4-6.5) |
| Regions |  |  |  |
| Guadeloupe | 0.3% (0.0-0.7) | 2.7% (1.4-4.4) | 3.3% (1.7-5.1) |
| Martinique | 0.2% (0.0-0.5) | 2.0% (1.0-3.4) | 2.4% (1.2-3.9) |
| French Guiana | 0.6% (0.1-1.4) | 6.0% (3.3-9.9) | 7.1% (4.0-11.5) |
| La Reunion | 0.3% (0.0-0.8) | 2.5% (0.8-4.9) | 3.0% (1.0-5.8) |
| Île-de-france | 0.8% (0.1-1.6) | 7.4% (5.7-9.4) | 8.8% (6.9-11.0) |
| Centre-Val-de-Loire | 0.3% (0.0-0.7) | 2.6% (1.1-4.6) | 3.1% (1.3-5.4) |
| Bourgogne-Franche Comté | 0.4% (0.0-0.9) | 3.6% (1.9-5.8) | 4.3% (2.2-6.9) |
| Normandie | 0.3% (0.0-0.8) | 3.3% (1.9-5.1) | 3.9% (2.3-6.0) |
| Hauts-de-France | 0.3% (0.0-0.7) | 2.9% (1.7-4.4) | 3.5% (2.1-5.3) |
| Grand-Est | 0.7% (0.1-1.7) | 7.2% (4.8-10.2) | 8.6% (5.8-11.8) |
| Pays de la Loire | 0.3% (0.0-0.7) | 2.9% (1.4-4.9) | 3.5% (1.8-5.9) |
| Bretagne | 0.2% (0.0-0.6) | 2.1% (0.8-4.0) | 2.6% (1.0-4.8) |
| Nouvelle-Aquitaine | 0.3% (0.0-0.6) | 2.6% (1.5-4.0) | 3.2% (1.8-4.7) |
| Occitanie | 0.3% (0.0-0.6) | 2.6% (1.3-4.1) | 3.1% (1.6-4.9) |
| Auvergne-Rhône-Alpes | 0.4% (0.0-0.8) | 3.7% (2.3-5.4) | 4.4% (2.8-6.5) |
| Provence-Alpes-Côte d'Azur | 0.3% (0.0-0.6) | 2.8% (1.6-4.2) | 3.3% (2.0-4.9) |
| Corse | 0.3% (0.0-0.8) | 3.2% (1.3-5.8) | 3.8% (1.5-6.9) |

Legend: Prevalence of SARS-CoV-2 antibodies was based on at least one positive test among LuLISA S, LuLISA N and pseudo-neutralisation assays (see main text).

**Figure S1:**
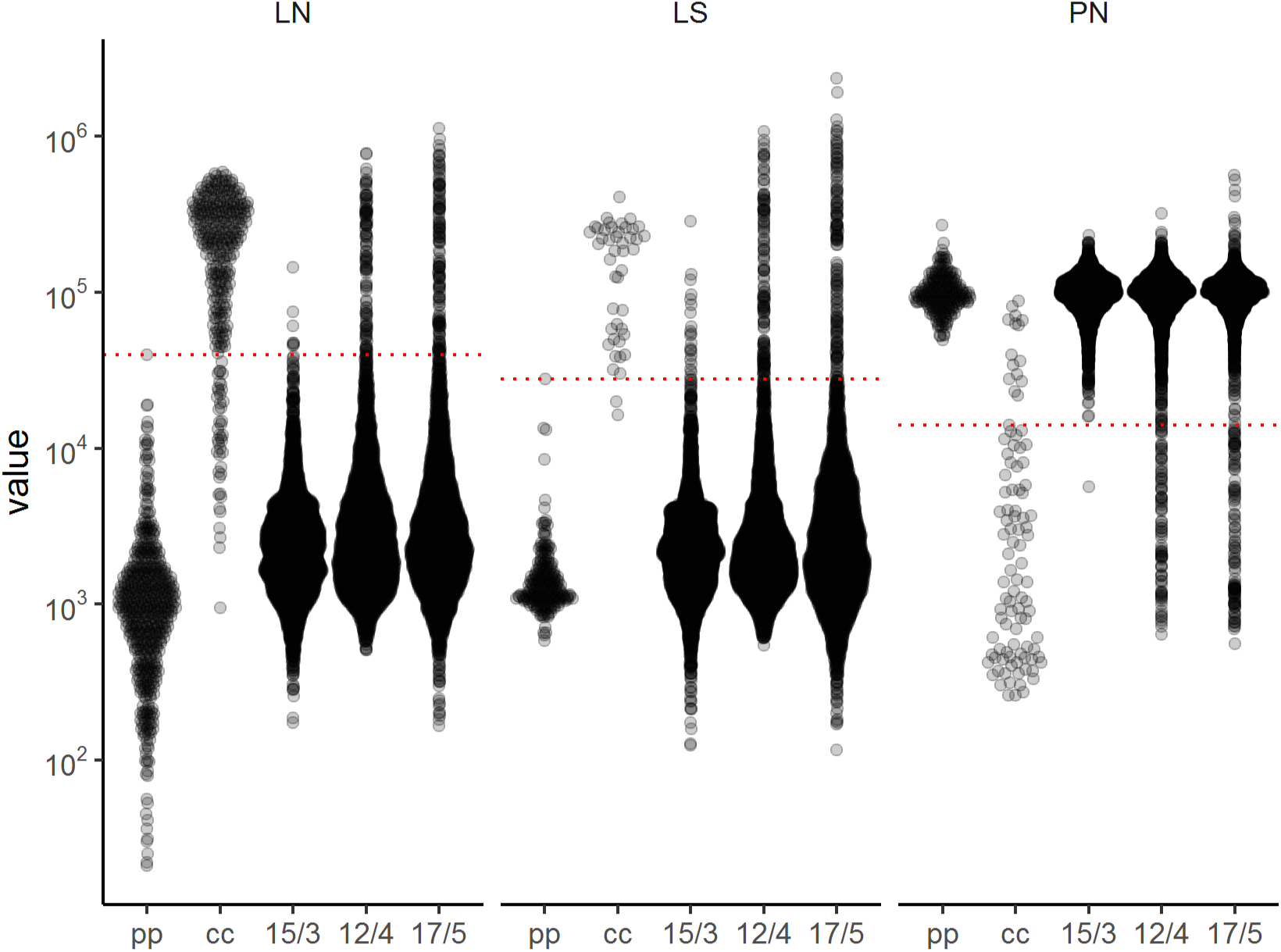
Distribution of quantitative values for the LuLISA N, LuLISA S and pseudo-neutralisation assays. Readings in relative light units (RLU in logarithmic scale) are presented LuLISA N (LN), LuLISA S (LS) and pseudo-neutralisation (PN) assays for sera from pre-pandemic (pp) patients, confirmed cases of COVID-19 (cc), and sera sampled during three collection periods 15/3 (9-15 March 2020), 12/4 (6-12 April 2020) and 17/5 (11-17 May 2020). Positivity thresholds are indicated by horizontal dotted lines.

**Figure S2:**
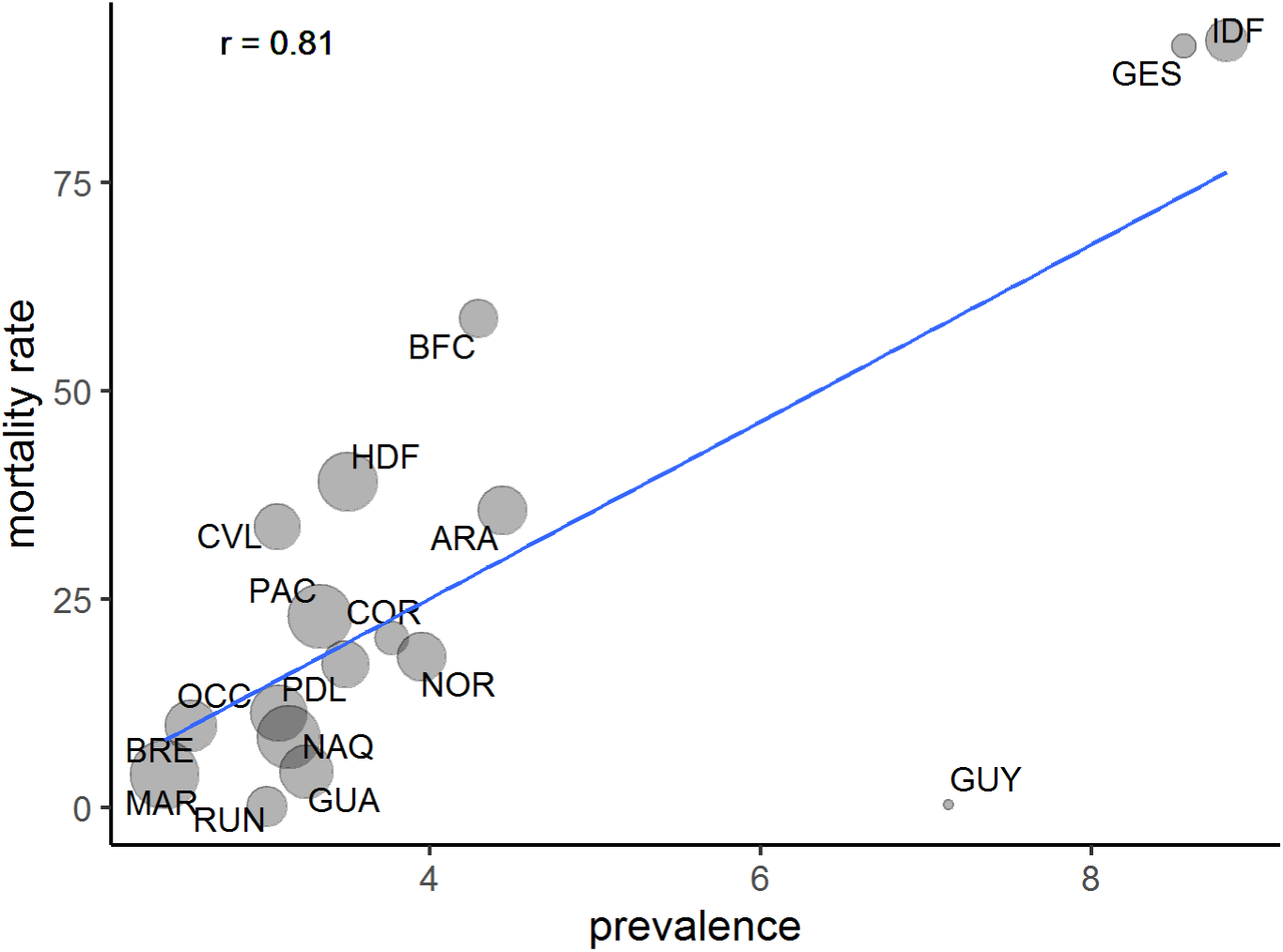
Weighted correlation between estimated prevalence of SARS-CoV-2 antibodies and reported mortality rates by region. Mortality rates per 100 000 were obtained as region-specific number of deaths attributed to COVID-19 as of 29 May 2020 divided by population size. The date to account for deaths was calculated assuming that individuals with detectable antibodies at sampling time (midpoint of interval from 11 to 17 May 2020) could have been infected at minimum 15 days previously and were susceptible of dying from their infection up to 30 days post-infection. Pearson correlation coefficient (r) was weighted by standard error of seroprevalence estimates. Circle sizes reflect this weighting. Regions: GUA = Guadeloupe, MAR = Martinique, GUY = French Guiana, RUN = La Réunion, IDF = Île-de-France, CVL = Centre-Val-de-Loire, BFC = Bourgogne-Franche Comté, NOR = Normandie, HDF = Hauts-de-France, GES = Grand-Est, PDL = Pays de la Loire, BRE = Bretagne, NAQ = Nouvelle-Aquitaine, OCC = Occitanie, ARA = Auvergne-Rhône-Alpes, PAC = Provence-Alpes-Côtes d’Azur, COR = Corse.

